# Life Satisfaction Among Cancer Survivors With and Without Smell and Taste Dysfunction: Evidence from the National Health Interview Survey

**DOI:** 10.1101/2025.11.03.25339420

**Authors:** Victoria Esparza, Vicente Ramirez, Alissa Nolden, Kara Stromberg, Valentina Parma

**Author notes:** Co-first authors (Victoria Esparza, Vicente Ramirez). Corresponding author: Valentina Parma.

## Abstract

**Background:** Chemosensory dysfunction is a common symptom experienced by cancer patients. Albeit common, these symptoms lack guidelines for management and are rarely addressed by healthcare providers, and therefore, how taste and smell dysfunctions affect quality of life in cancer patients is understudied.

**Methods:** To address this gap, we analyzed data from the 2021 National Health Interview Survey (NHIS) to assess associations between smell and taste dysfunction and life satisfaction among adults who self-reported cancer (N = 3,327; age range:18- 85+ years; 57% F; 85% Non-Hispanic white). Participants reported whether they experienced difficulty tasting, difficulty smelling, and whether they were satisfied with life (binary) in the past 12 months.

**Results:** Among respondents, 14% reported taste dysfunction (N = 456) and 22% reported smell dysfunction (N = 736), and 10.1% reported both smell and taste dysfunction (N = 340). After adjusting for age and sex, a logistic regression revealed that cancer patients reporting difficulty tasting had 60% lower odds of reporting life satisfaction compared to those who did not report difficulty tasting (OR = 0.39, p<0.0001, CI [.27,.58]). Similarly, cancer patients reporting difficulty smelling had 40% lower odds of reporting life satisfaction compared to those who did not report trouble smelling (OR = 0.60, p=0.006, CI [.42,.86]). Considering that the prevalence of chemosensory dysfunction is often underestimated when measured with self-reports, these findings likely represent a conservative estimate of the impact on subjective wellbeing in cancer.

**Conclusion:** These results underscore the relevance of assessing chemosensory symptoms in cancer populations. Routine evaluation of smell and taste loss may offer a valuable opportunity to improve quality of life and guide supportive care.

## Background

Chemosensory dysfunction, namely the loss or alteration of smell (olfaction) and taste (gustation), is an important but often overlooked consequence of cancer and its treatment (1). Recently, smell and taste disorders have gained wider attention due to their prominence in COVID-19 (affecting roughly 60% of COVID patients) (2). However, such disorders are not unique to viral infections; they are also frequent in several inflammation-based conditions (3), as well as in oncology. Taste and smell abnormalities are common in people with cancer, yet historically they have been ignored or underestimated in this population (4). Studies indicate that approximately one-fifth to over half of patients experience altered taste during cancer therapies (4), and a significant fraction report impaired smell (5). In women with breast cancer undergoing chemotherapy, for example, taste changes have been reported in about 56– 76%, while 16–49% report smell changes (6). Some reports even suggest that up to 93% of patients across various cancers complain of taste disturbance and around 60% report changes in smell (1). Notably, these chemosensory side effects can span the entire disease course and may persist well into survivorship, long after treatment has concluded (7). In other words, a subset of cancer survivors continues to experience chronic smell or taste loss as a lasting legacy of their illness or therapy.

These chemosensory deficits have significant implications for patients’ health-related quality of life and life satisfaction. The senses of taste and smell are intimately tied to the enjoyment of food and beverages, appetite regulation, and social customs like shared meals (8,9). When taste or smell is blunted or distorted, patients may develop food aversions or a lack of interest in eating, leading to poor nutritional intake, unintentional weight loss, and low energy (10–12). Indeed, taste/smell alterations are linked to an increased risk of malnutrition and can contribute to cancer-associated anorexia–cachexia syndrome (13).

Beyond nutrition, patients often experience psychological and social consequences: many report that distorted senses of taste/smell dampen their mood and enjoyment of life, and they may withdraw from social situations centered around food (14). Studies have documented that chemosensory changes correlate with worse overall quality of life in cancer patients (14).

In short, what might seem like “minor” sensory nuisances can cascade into broader physical and emotional challenges, undermining both day-to-day happiness and clinical outcomes. For instance, untreated taste and smell problems can reduce oral intake, weakening immunity and even hindering adherence to treatments (12). Patients with chemosensory dysfunction have increased risk of malnutrition and associated decreased clinical outcomes (15). Malnutrition impacts upwards of 80% of cancer patients at some point during their cancer journey. Weight loss of 6% predicts a reduced response to treatment, reduced survival, and reduced quality of life (16). Ultimately, patients with unresolved chemosensory dysfunction may experience reduced life satisfaction as they struggle with the loss of fundamental pleasures and normalcy in daily living.

Despite their prevalence and impact, smell and taste disturbances remain under- recognized in cancer care. Compared to more visible side effects (such as nausea or pain), clinicians often give less attention to chemosensory complaints. In practice, changes in taste and smell changes are rarely assessed or documented during oncology visits (17). There are currently no clinical guidelines or standardized protocols to screen for or manage cancer-related taste and smell disorders (1). Many oncology providers have not been formally trained to address these symptoms, and patients are seldom proactively informed about the possibility of developing chronic smell/taste loss. This gap in clinical guidance means that interventions (i.e., nutritional counseling, olfactory training, flavor enhancement strategies, etc.) are not routinely offered, leaving patients to cope on their own. The under-recognition of chemosensory dysfunction in survivorship care plans represents a missed opportunity to improve patients’ quality of life (17). In light of increasing cancer survival rates and emphasis on survivorship well-being, it is crucial for the medical community to reframe taste and smell not as trivial issues but as critical facets of patient care.

To date, most research on cancer-related taste and smell changes has been limited to specific clinics or subsets of patients (for example, trials in head and neck cancer or small cohort studies in chemotherapy settings) (7, 18). Consequently, little is known about the broader, population-level impact of chemosensory dysfunction on survivors’ lives. In particular, the relationship between these sensory deficits and survivors’ overall subjective well-being remains insufficiently explored. Population-based data can add valuable insights here: by capturing diverse cancer survivors in the community, such data allow us to examine associations between self-reported taste/smell dysfunction and outcomes like life satisfaction on a large scale. Life satisfaction is a key indicator of subjective well-being, reflecting an individual’s overall appraisal of their life quality and happiness. Understanding whether survivors with chemosensory problems have diminished life satisfaction can highlight the real-world significance of these symptoms beyond clinical metrics. However, to our knowledge, no prior study has assessed this association in a nationally representative sample of survivors.

The present study addresses a specific gap: using data from the National Health Interview Survey (NHIS), a large population-based health survey, we investigate the association between self-reported chemosensory dysfunction and life satisfaction in U.S. adults with a history of cancer. By leveraging recent NHIS data, we aim to quantify how loss of smell and/or taste correlates with an individual’s satisfaction with life, after accounting for possible confounding factors, such as age and sex. We expect cancer survivors suffering from chemosensory impairments to report lower subjective well-being compared to their peers without such impairments, revealing the need for consideration of these symptoms in oncological and survivorship care, ultimately contributing to better health-related quality of life outcomes for the growing population of cancer survivors.

## Methods

### Data Source and Sample Population

This study utilized data from the National Health Interview Survey (NHIS) which is a nationally representative cross-sectional survey conducted annually by the National Center for Health Statistics (NCHS). To examine life satisfaction among cancer patients, we analyzed the adult data from the 2021 Sample Adult questionnaire which includes variables on self-assessed taste and smell function, cancer history, and self-evaluation of life-satisfaction and well-being. We restricted the analytic sample to adult respondents (aged ≥18) who self-reported if they have ever been diagnosed with cancer (can_ev). Respondents with missing data on key variables (e.g., self-reported chemosensory function, life satisfaction, age, sex, race, health insurance coverage, and urban-rural classification) because they were not assessed, refused to report, or did not know how to report were excluded from the primary analysis. We analyzed adults (18- 85+) with a self-reported history of cancer (n = 3,327). The majority of respondents were aged 66-84 (55%), female (57%), and non-Hispanic White (85%). In the unweighted sample, approximately 94% of participants reported being satisfied with life, while 6% reported dissatisfaction. Among cancer survivors in the sample around 14% reported some form of taste dysfunction (either current difficulty or worse than past) and around 22% reported smell dysfunction. Please refer to Table 1 for details.

**Table 1.**
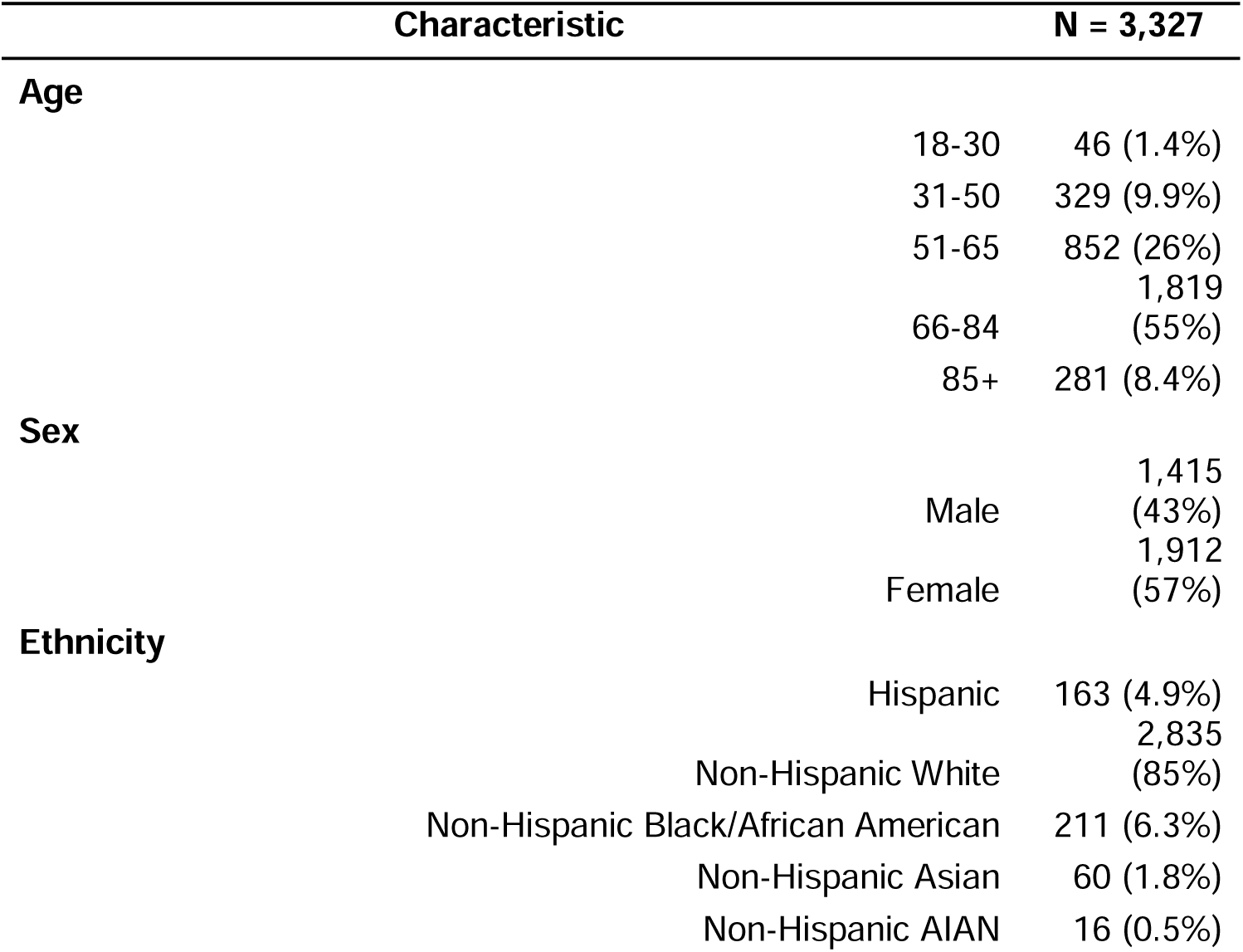
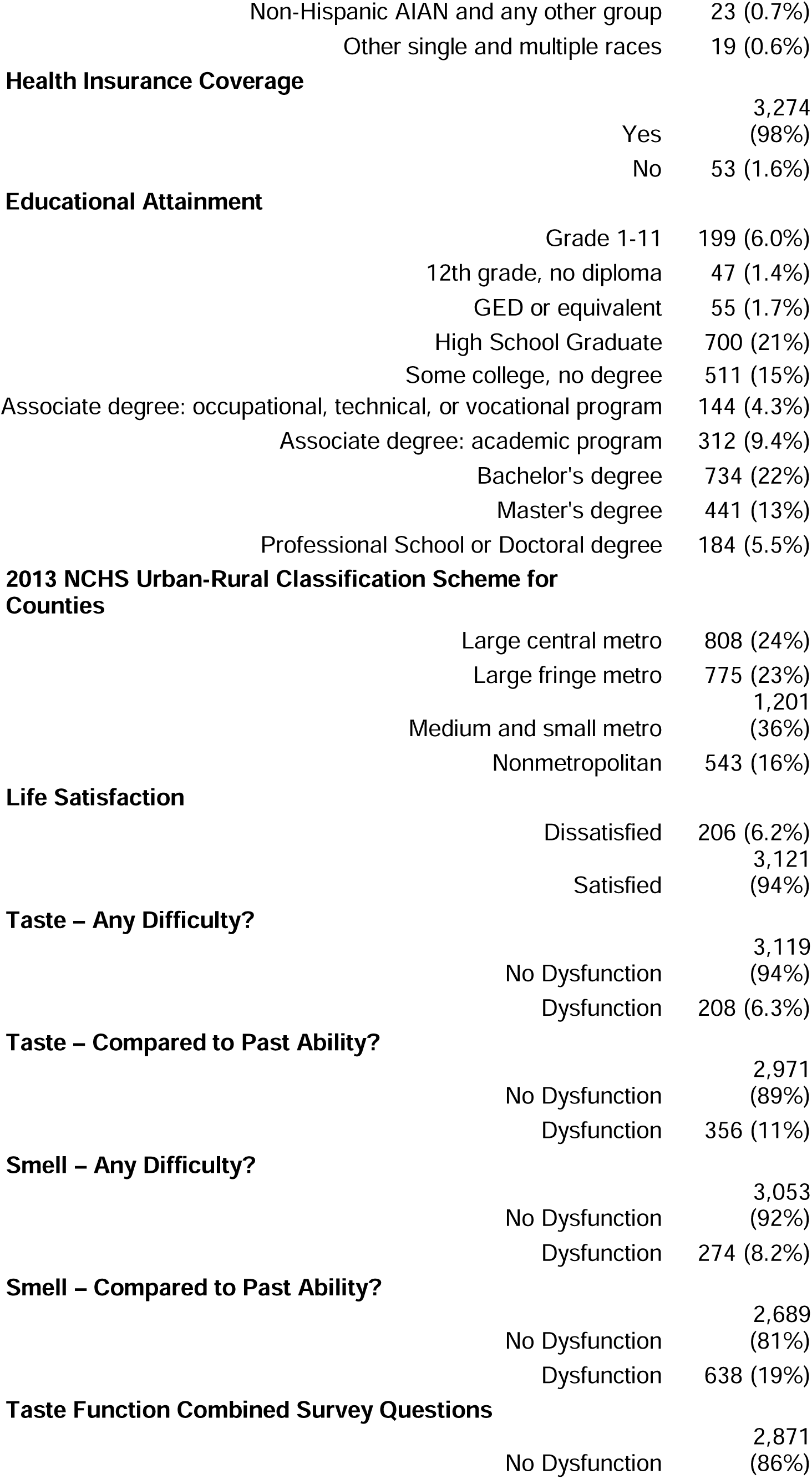
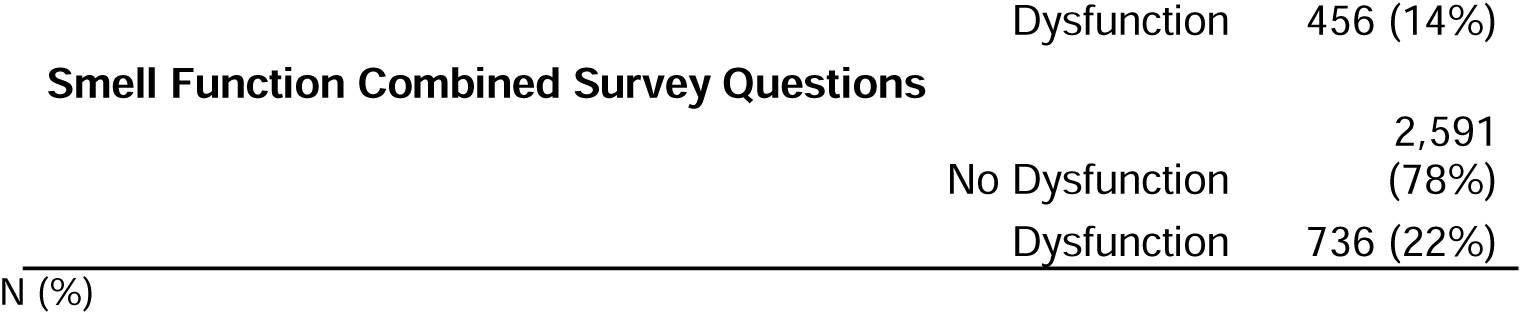
Demographic characteristics of participants included in the present study. .

### Measures

#### Data Transformation and Survey Design

To generate nationally representative estimates and preserve the validity of standard errors, all analyses accounted for the complex survey design of NHIS by incorporating strata (*pstrat*), primary sampling units (*ppsu*), and sample adult weights (*wtfa_a*). Survey design objects were created using the *survey* and *srvyr* packages in R (14)(19). The survey design was defined prior to filtering and transforming data. The final analytic dataset was restricted to individuals with complete data for cancer history, chemosensory function, life satisfaction, age (*agep_a*), sex (*sex_a*), race/ethnicity (*hispallp_a*), educational attainment (*educp_a),* insurance coverage (*hicov_a*), and urban-rural classification (*urbrrl*). All observations that had missing data in observations were removed from these variables of interest. In the NHIS, age is top-coded at 85 years old, which necessitated creating aggregating age groups into the following bins: 18-30, 31-50, 51-65, 66-84, and 85+.

Life satisfaction was assessed as a 4-level item (*lsatis4r_a*) and dichotomized as satisfied (“very satisfied“or”satisfied") and dissatisfied (“dissatisfied” or “very dissatisfied”). Self-reported chemosensory function was derived from questions related to smell and taste in the NHIS questionnaire. For smell, we utilized two NHIS variables: smellcomp_a (change in ability compared to past) and smelldf_a (current difficulty smelling). For taste, we utilized tastecomp_a (change in ability compared to past) and tastedf_a (current difficulty tasting). Chemosensory dysfunction, for smell and taste assessed separately, was defined based on three criteria: perceived change from past function, self-reported difficulty, or a combined indicator, in which individuals were classified as having dysfunction if they endorsed either current difficulty or worsening of one chemical sense.

### Statistical Analysis

Survey-weighted binary logistic regression models were used to estimate the association between chemosensory dysfunction and life satisfaction (dichotomized as satisfied vs. not satisfied). For each modality (taste and smell), we constructed separate models using each criterion (self-reported difficulty, perceived change from past function, or a combined indicator). All models were adjusted for age and sex, as these variables are highly associated with chemosensory perception and function. Regression models were further adjusted for race (raceallp_a), educational attainment (educp_a), healthcare coverage (cover_a), and urban-rural classification (urbrrl). All analyses and visualization of these results were carried out in *R* version 4.3.3. Forest plots depicting the exponentiated model coefficients (odds ratios) were generated using the *ggplot2* package. P-values were calculated using the Wald approximation and then adjusted for multiple comparisons using the Benjamini-Hochberg method.

## Results

### Chemosensory Dysfunction and Life Satisfaction

After controlling for the effects of age and sex in the model, taste dysfunction was significantly associated with decreased odds of life satisfaction (**Figure 1**). Participants reporting their taste was worse than in the past had on average 66% lower odds of reporting positive life-satisfaction (OR = 0.34, 95% CI: 0.22-0.51, p < 0.001). Those reporting current taste difficulty had 50% lower odds (OR = 0.50, 95% CI: 0.30-0.825, p = 0.007). The combined indicator of taste dysfunction has a 60% reduction in the odds of reporting life-satisfaction (OR=0.39, 95% CI: 0.27-0.58, p < 0.001).

**Figure 1.**
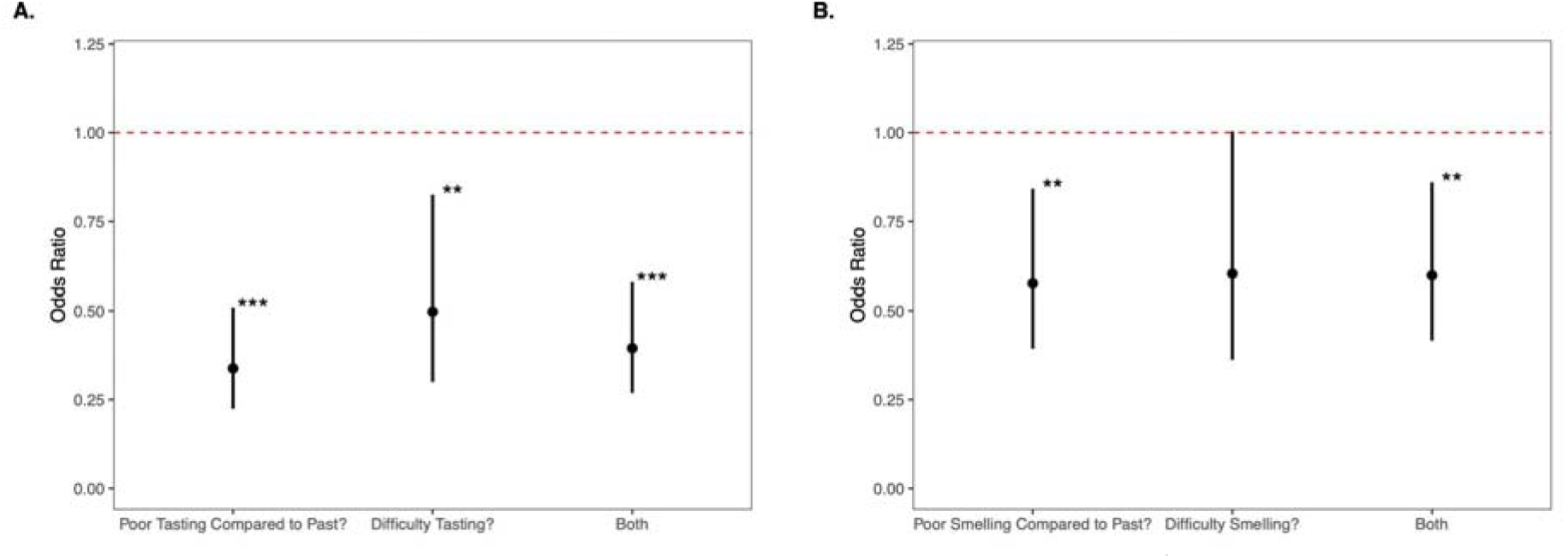
Unadjusted odds ratios for reported life satisfaction and taste and smell dysfunction. Odds ratio of decreased life satisfaction for participants with cancer and taste and/or smell dysfunction after controlling for sex and age. Black dots represent the odds ratio, and bars represent the 95% confidence interval. The dotted red line represents the baseline odds at OR =1 * p>0.05, ** p>0.01, *** p>0.001

Smell dysfunction was also independently associated with reduction in the odds of life satisfaction (**Figure 1**). We found that those that reported that their sense of smell is worse than in the past on average had 42% reduced odds of reporting positive life satisfaction (OR = 0.58, 95% CI: 0.39-0.84, p = 0.005). Those that reported current smell difficulty were found to have a 40% reduction in the odds of reporting positive life satisfaction; however, this finding showed a tendency but did not meet the threshold for statistical significance (OR = 0.60, 95% CI: 0.36-1.00, p = 0.052). When a combined definition of smell dysfunction is used, smell dysfunction is associated with a 40% reduction in the odds of reporting life-satisfaction (OR = 0.60, 95% CI: 0.42-0.86, p = 0.006).

After further adjusting the model to control for the effects of race, educational attainment, insurance coverage, and urban-rural classification, the association between taste dysfunction and life dissatisfaction remained significant (**Figure 2**). The adjusted model found that those who reported worse taste compared to the past on average had a 68% reduction in the odds of reporting positive life satisfaction (OR = 0.32, 95% CI: 0.21-0.49, p<0.001) and when we examined those that reported a current difficulty in tasting we found a 48% reduction in the odds of positive life-satisfaction (OR = 0.52, 95% CI: 0.31-0.87, p = 0.013). The combined definition for taste dysfunction yielded a 61% reduction in these odds (OR = 0.39, 95% CI: 0.26-0.57, p< 0.001).

**Figure 2.**
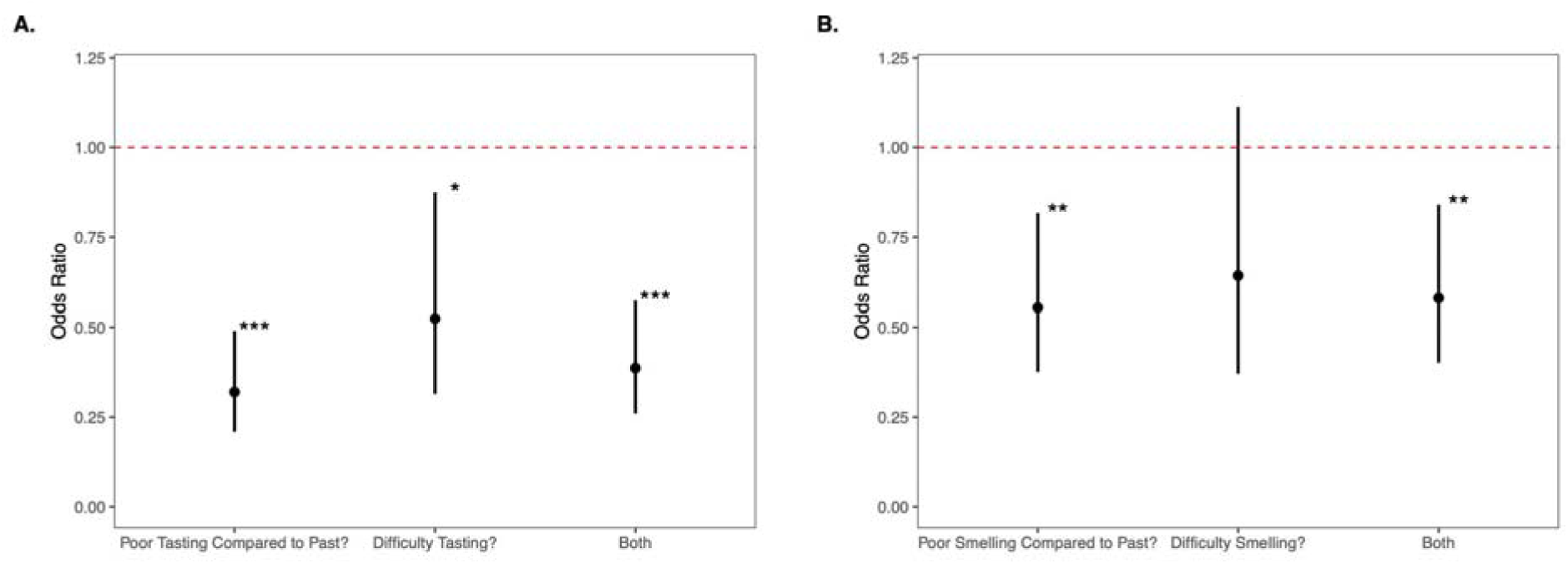
Adjusted odds ratios for reported life satisfaction and taste and smell dysfunction. Odds ratio of decreased life satisfaction for participants with cancer and taste and/or smell dysfunction after controlling for age, sex, race, educational attainment, insurance coverage, and urban-rural classification. Black dots represent the odds ratio, and bars represent the 95% confidence interval. The dotted red line represents the baseline odds at OR=1. * p>0.05, ** p>0.01, *** p>0.001

Similarly, after adjustment the effect of smell dysfunction on life satisfaction remained significant, but not across all metrics. After adjusting the model, we found that those individuals who reported that their sense of smell was worse than in the past had 45% decreased odds of reporting life satisfaction (OR = 0.55, 95% CI: 0.38-0.82, p = 0.003). The association between life satisfaction and current difficulty smelling resulted in a 36% reduction in the odds of reporting positive life-satisfaction, but this was not significant (OR = 0.64, 95% CI: 0.39-1.1, p = 0.114). The combined definition remained significant, yielding a 42% reduction in the odds of reporting positive life satisfaction (OR = 0.58, 95% CI: 0.40-0.84, p = 0.004).

We additionally assessed the association between life satisfaction and sensory dysfunction when both smell and taste dysfunction are simultaneously reported. As indicated in **Supplementary Table 1** the reduction in life satisfaction is greater (OR = 0.35, 95% CI: 0.22-0.54, p < 0.001) in those that report both smell and taste dysfunction.

## Discussion

In this study, we found that self-report of smell and taste dysfunction is a potential predictor of life satisfaction in individuals with a history of cancer. The findings showed individuals with a history of cancer who self-report having difficulties in taste and smell function had 40-61% lower odds of reporting positive life satisfaction in comparison to their peers who do not have such impairments. These associations remained significant after adjustment of our model for social, environmental, and demographic factors, suggesting that the link between chemosensory dysfunction and reduced life satisfaction in cancer patients is independent of these factors. This result supports the view that the impact of chemosensory dysfunction on life satisfaction is not merely dependent on individual or environmental context but is instead intrinsically tied to the sensory loss itself.

### Taste dysfunction is strongly associated with life dissatisfaction

While unmeasured in our current study, a number of potential mechanisms might cause taste dysfunction among cancer patients. Tumor location, especially in head and neck, cancers, and thoracic cancers such as esophageal cancer, may contribute directly to the taste dysfunction seen among patients (20). Chemotherapy and radiation therapy may cause biological and physiological changes in salivary gland composition, hyposalivation, mucosal alterations, epithelial tissue damage, and neurotoxicity which might results in taste and smell alterations or impairment of chemosensory systems (20). Overall, the mechanisms that drive and treat cancer may cause changes in both taste and smell perception. Interestingly, these findings are stronger in those who report taste dysfunction compared to those reporting smell dysfunction. While both gustation and olfaction play vital roles in patients’ health-related quality of life and life satisfaction, it appears that the effects on taste are most prominent. These findings are consistent with prior meta-analysis work conducted by Nolden and colleagues (2019), who found that taste alterations were reported more frequently than smell amongst cancer patients. However, when we examined the effects of simultaneous smell and taste dysfunction, we found that the association between life satisfaction and sensory dysfunction is most pronounced in those that report both smell and taste dysfunction (**Supplementary Table 1**). Due to the self-reported nature of the data (NHIS 2021) and known confusion between chemosensory modalities in self-assessment (21,22), these findings should be interpreted with caution. For example, patients frequently misattribute retronasal olfactory deficits (critical for flavor perception) as problems with their taste (23,24).

During the COVID-19 pandemic, meta-analyses revealed that self-report underestimates olfactory dysfunction by up to 30% compared to psychophysical testing (25). In contrast, a similar discrepancy was not observed for taste dysfunction, perhaps due to the limited availability and lower standardization of psychophysical gustatory assessments (26). Taken together, it is plausible that some individuals reporting taste dysfunction may, in fact, be experiencing unrecognized olfactory deficits.

### Chemosensory Dysfunction Suggest Unique Vulnerabilities for Cancer Patients

These results suggest that cancer patients that acquire taste and smell loss may face unique vulnerabilities that may result in decrements in their quality of life and overall life satisfaction. Chemosensory dysfunction is associated with deterioration of nutritional status, which may further exacerbate the compounded effect of cancer on patients and lead to increased risk of malnutrition. In patients undergoing chemotherapeutic intervention that report chemosensory dysfunction, there is an increased risk of non-compliance in clinically directed use of oral nutritional supplements to prevent nutritional deficiencies and malnutrition (27). Further, chemosensory dysfunction is associated with psychological and emotional distress (14), and its presence in cancer patients may indicate increased risk of exacerbated stressors in patients. In these contexts, the presence of chemosensory function may act as a predictor of physiological and psychological morbidities, quality of life, and mortality in cancer patients.

### Clinical Implications

Olfactory and gustatory function is underassessed in the clinical setting and has historically been ignored as a symptom of interest in oncological care (17). The association between decreased life satisfaction and chemosensory dysfunction suggests that clinicians may be able to improve patients’ quality of life through targeted screening of chemosensory disorders in cancer patients. These results suggest that clinicians, therapists, and the patient support team may benefit from integrating psychophysical chemosensory tools and structured patient questionnaires aimed at addressing chemosensory deficits throughout the cancer continuum. Such screenings could inform and guide supportive patient interventions, including psychological counseling and mental health support, nutritional counseling, and guide symptom monitoring strategies.

However, to date, there are no standardized guidelines for screening or addressing decrements in chemosensory dysfunction in cancer patients. Recently developed taste and smell tests that offer rapid protocols show potential deployment in healthcare settings (28–30); however, their adoption in health care settings and in oncological care is not widespread. The broader implementation of such tools could potentially fill an important gap in survivorship care for cancer patients and may offer new, holistic, and patient-centered approaches for preserving quality of life.

### Study Limitations

The current study is not without limitations. The cross-sectional nature of the NHIS data does not allow for inferences to be drawn on the temporality of the elucidated association (e.g., if chemosensory dysfunction preceded changes in life satisfaction), and therefore any speculation on causal inferences from our results. Additionally, limited information on the timeline of cancer diagnosis, the onset of chemosensory deficits, and the timeline of therapeutic intervention is limited in this data. The data are also limited in the detail of the clinical context for each patient, and do not shed light on treatment history, cancer staging and severity, and other relevant clinical details. As addressed previously, the assessment of chemosensory function in the study is based on self-report, which is prone to measurement error through both recall bias and misclassification of reported symptoms through confusion of taste and retronasal olfaction.

Regarding the outcome of interest, the life satisfaction measurement is limited in that it does not provide domain-specific dimensions of life satisfaction, and therefore it is likely vulnerable to unmeasured random effects and potential confounders. Single-item measures limit reliability and tend to be sensitive to transient or situational influences on responses. Dimensions of life satisfaction and quality of life such as food enjoyment, social connectedness, autonomy, and existential fulfillment are among the different aspects of life satisfaction that may be relevant to the variation in the single measurement used in this study, that are otherwise aggregated into a single measure. Therefore, our results should be interpreted cautiously, and future studies should seek to use validated and multidimensional methods of measurement for both chemosensory function and life satisfaction.

## Conclusion

The present study suggests that chemosensory dysfunction has a moderate influence on life satisfaction in cancer patients. Our findings reveal that cancer survivors suffering from chemosensory impairments have higher odds of reporting lower life satisfaction in comparison to their peers without such impairments. Therefore, the current study suggests that chemosensory function is a highly relevant and actionable, though often overlooked, aspect of supportive care in cancer patients. Implementing chemosensory tools in patient care may offer new avenues for preserving life satisfaction and quality of life among cancer patients.

## Authors contributions

Based on Credit taxonomy, V.E., V.R., V.P. developed the research question and overall study framework, V.R., V.P. designed the analytic approach and selected relevant variables from NHIS, V.R. curated the data, conducted the analyses and created visualizations, V.E., V.R., V.P. drafted the initial manuscript, V.E., V.R., A.N., K.S., V.P. revised the manuscript critically for important intellectual content, V.P. oversaw the project.

## Ethics approval

This study was based on publicly available, de-identified data from the U.S. National Health Interview Survey (NHIS), which is conducted annually by the National Center for Health Statistics (NCHS). The NHIS is approved by the Research Ethics Review Board of the National Center for Health Statistics, Centers for Disease Control and Prevention. No additional ethical approval from an Institutional Review Board (IRB) was required for this secondary data analysis.

## Consent to participate

Informed consent was obtained from all individuals included in the study.

## Consent for Publication

The requirement for consent to publish is not applicable for this study.

## Availability of data and materials

The datasets generated and analyzed during the current study are available in the CDC National Center For Health Statistics repository,(31)

## Data Availability

All datasets analyzed in the current study are available in the CDC National Center For Health Statistics repository. All data generated in this study are available upon request.

https://www.cdc.gov/nchs/nhis/documentation/2021-nhis.html

## Acknowledgments

We thank the National Center for Health Statistics and the survey participants who contributed to the NHIS.

