## Supplementary Table 1 for "Life Satisfaction Among Cancer Survivors With and Without Smell and Taste Dysfunction: Evidence from the National Health Interview Survey"

| **Characteristic** | **OR** | **95% CI** | **p-value** |
| --- | --- | --- | --- |
| **Age** |  |  |  |
| 18-30 | — | — |  |
| 31-50 | 1.25 | 0.46, 3.39 | 0.7 |
| 51-65 | 1.44 | 0.56, 3.66 | 0.4 |
| 66-84 | 2.36 | 0.92, 6.03 | 0.073 |
| 85+ | 1.70 | 0.59, 4.91 | 0.3 |
| **Sex** |  |  |  |
| Male | — | — |  |
| Female | 1.04 | 0.74, 1.47 | 0.8 |
| **Ethnicity** |  |  |  |
| Hispanic | — | — |  |
| Non-Hispanic White only | 0.80 | 0.37, 1.74 | 0.6 |
| Non-Hispanic Black/African American only | 0.47 | 0.19, 1.14 | 0.093 |
| Non-Hispanic Asian only | 0.81 | 0.21, 3.15 | 0.8 |
| Non-Hispanic AIAN only | 0.29 | 0.06, 1.39 | 0.12 |
| Non-Hispanic AIAN and any other group | 8.82 | 0.92, 84.8 | 0.060 |
| Other single and multiple races | 0.16 | 0.02, 1.09 | 0.062 |
| **Health Insurance Coverage** |  |  |  |
| Yes | — | — |  |
| No | 0.41 | 0.18, 0.93 | 0.034 |
| **Educational Attainment** | | |  |
| Grade 1-11 | — | — |  |
| 12th grade, no diploma | 1.13 | 0.29, 4.45 | 0.9 |
| GED or equivalent | 0.56 | 0.21, 1.53 | 0.3 |
| High School Graduate | 1.36 | 0.70, 2.66 | 0.4 |
| Some college, no degree | 1.83 | 0.91, 3.68 | 0.092 |
| Associate degree: occupational, technical, or vocational program | 1.46 | 0.55, 3.90 | 0.4 |
| Associate degree: academic program | 1.69 | 0.79, 3.63 | 0.2 |
| Bachelor's degree | 2.65 | 1.29, 5.43 | 0.008 |
| Master's degree | 6.15 | 2.32, 16.3 | <0.001 |
| Professional School or Doctoral degree | 3.30 | 1.19, 9.14 | 0.022 |
| **2013 NCHS Urban-Rural Classification Scheme for Counties** | | | |
| Large central metro | — | — |  |
| Large fringe metro | 1.22 | 0.75, 2.00 | 0.4 |
| Medium and small metro | 1.27 | 0.82, 1.96 | 0.3 |
| Nonmetropolitan | 1.43 | 0.82, 2.52 | 0.2 |
| **Combined Taste and Smell Dysfunction Reported** | |  |  |
| No Dysfunction | — | — |  |
| Smell only | 0.95 | 0.53, 1.70 | 0.9 |
| Taste only | 0.51 | 0.25, 1.08 | 0.078 |
| Both | 0.35 | 0.22, 0.54 | <0.001 |
